# Estimated Impacts of Rotavirus Vaccine Recommendation Changes in the U.S.

**DOI:** 10.64898/2026.04.27.26351857

**Authors:** Ernest O. Asare, Jiye Kwon, Melanie H. Chitwood, Stephanie Perniciaro, Gregg S. Gonsalves, Virginia E. Pitzer

## Abstract

In January 2026, the United States Department of Health and Human Services downgraded the recommendation for infant immunization with rotavirus vaccine to one of shared clinical decision-making. We use a validated model for the transmission dynamics of rotavirus to predict the magnitude and timing of increases in the number of rotavirus hospitalizations in the US and in representative states given possible decreases in vaccine coverage. Rotavirus hospitalizations are likely to increase within two to three years following a drop in vaccine coverage, resulting in over 200,000 hospitalizations between July 2026-June 2031 if coverage were to drop to 20%. The burden is likely to fall disproportionately on southern states that currently experience higher rates of rotavirus hospitalization.

## Main Text

Rotavirus is a leading cause of acute gastroenteritis in children worldwide. Nearly every child is infected with rotavirus by 3 to 5 years of age, potentially causing severe watery diarrhea, vomiting, fever and stomach pain in infants without preexisting immunity (*1*). Severe rotavirus-associated gastroenteritis (RVGE) can lead to dehydration requiring hospitalization and, in rare cases, death. While there is no specific treatment for the disease, most patients in high-income countries receive oral or intravenous rehydration and recover. Before the introduction of vaccines in 2006, RVGE caused over 400,000 physician visits, 200,000 emergency department visits, and 55,000 hospitalizations annually in the United States (US) at an estimated cost of $1 billion (*2*).

Since the licensure of the three-dose RotaTeq vaccine in 2006 and two-dose Rotarix vaccine in 2008, there have been substantial and sustained declines in RVGE in the US (*3, 4*). Rotavirus vaccine effectiveness has been estimated to be 75% against emergency department visits and 82% against hospitalization based on data from the New Vaccine Surveillance Network (*5*); children with RVGE are four times more likely to require hospitalization if they have not been vaccinated against rotavirus (*5*). Vaccination averted an estimated 382,000 hospitalizations and $1.2 billion in medical costs between 2008 and 2013 (*6*) and notably changed the epidemiology of RVGE in the US, shifting from large annual outbreaks to smaller biennial outbreaks (*4*), as previously predicted by mathematical models (*7*).

Despite this success, the US Department of Health and Human Services (HHS) published guidelines in January 2026 downgrading the previous universal recommendation for infant rotavirus immunization to one that is dependent on shared clinical decision-making (SCDM), which requires that healthcare providers and caregivers first engage in a conversation about whether to vaccinate before the vaccine is prescribed. This recommendation was subsequently adopted by the Centers for Disease Control and Prevention (CDC). However, the American Academy of Pediatrics (AAP) continues to recommend the rotavirus vaccine for all infants without a history of intussusception, a rare but serious complication in which the intestine folds in on itself. AAP and other organizations have brought suit asking the court to enjoin the implementation of the January 2026 CDC guidelines, and a preliminary injunction was granted by the court in March 2026. Nonetheless, HHS has signaled that it will appeal the ruling, and thus the state of childhood immunization policy in the US remains in flux.

SCDM creates challenges to vaccine access. First, it presupposes that all families have access to healthcare and relationships with pediatricians or other healthcare professionals. This creates barriers for those who are un-or underinsured or who may live in rural or other areas with limited access to healthcare facilities (*8*). While HHS has suggested that all insurers should continue to cover vaccines under SCDM, it is unclear if insurance companies have the statutory obligation to do so or if HHS will continue to recommend coverage in the future (*9*). In addition, the costs for healthcare consultations may not be covered, requiring out-of-pocket expenditures for families. Finally, survey data indicates that recommendations for SCDM create confusion for providers and patients alike (*10*), including creating the perception that those vaccines are non-essential and of secondary clinical importance. This perception may be responsible for the fact that uptake of vaccines under SCDM (e.g. vaccines against meningococcal disease caused by serogroup B) lag far behind those under routine immunization categories (*11*). Thus, changes to rotavirus vaccine recommendations may lead to lower vaccine uptake and a resurgence of RVGE.

To predict the potential timing and magnitude of the resurgence in RVGE, we adapt a previously validated transmission dynamic model (*7*) for rotavirus to project RVGE hospitalizations in children under five. Reflecting current understanding of rotavirus epidemiology, we allow for re-infection at a lower rate after a period of temporary immunity and assume that the transmission rate varies seasonally. We model scenarios where vaccine coverage drops from current levels by 50% (relative decrease) or to 60%, 40%, or 20% coverage beginning in July 2026, reporting on outcomes through June 2031. We compare this to a base-case scenario where vaccine coverage is held constant at most recent reported levels. We calibrate our model to vaccine uptake data from the National Immunization Survey-Child (*12*) (**Supplemental Table S1**) and rotavirus hospitalization data (based on ICD-10CM code A08.0) from EPIC Cosmos (*13*) (**Supplemental Table S2**). To understand the impact of recommendation changes on specific states, we also use models fitted to data from California, Illinois, Massachusetts, Mississippi and Texas. These states differ in their current vaccine coverage, attitudes towards routine childhood vaccination (*14*), and birth rates (which impact rotavirus disease dynamics (*7*)). Additional model details can be found in the Supplementary Materials.

Based on the most recent available state-level estimates (2021 birth cohort), national coverage with a full rotavirus vaccine series was 75.4%, varying from 57.8% in Mississippi to 94.0% in Massachusetts. In the 2023-2025 rotavirus seasons, there were on average 7,660 hospitalizations annually in children <5 years old. If vaccine coverage amongst infants were to decline beginning in July 2026, we predict that RVGE hospitalizations would rebound within 2-3 years. Under the base-case scenario, we project 36,000 RVGE hospitalizations in the five-year period from July 2026 to June 2031, compared to 85,200 hospitalizations (2.4-fold increase) if vaccine coverage decreases modestly to 60% and 204,000 hospitalizations (5.7-fold increase) if vaccine coverage decreases dramatically to 20% (**Figure 1**).

**Figure 1.**
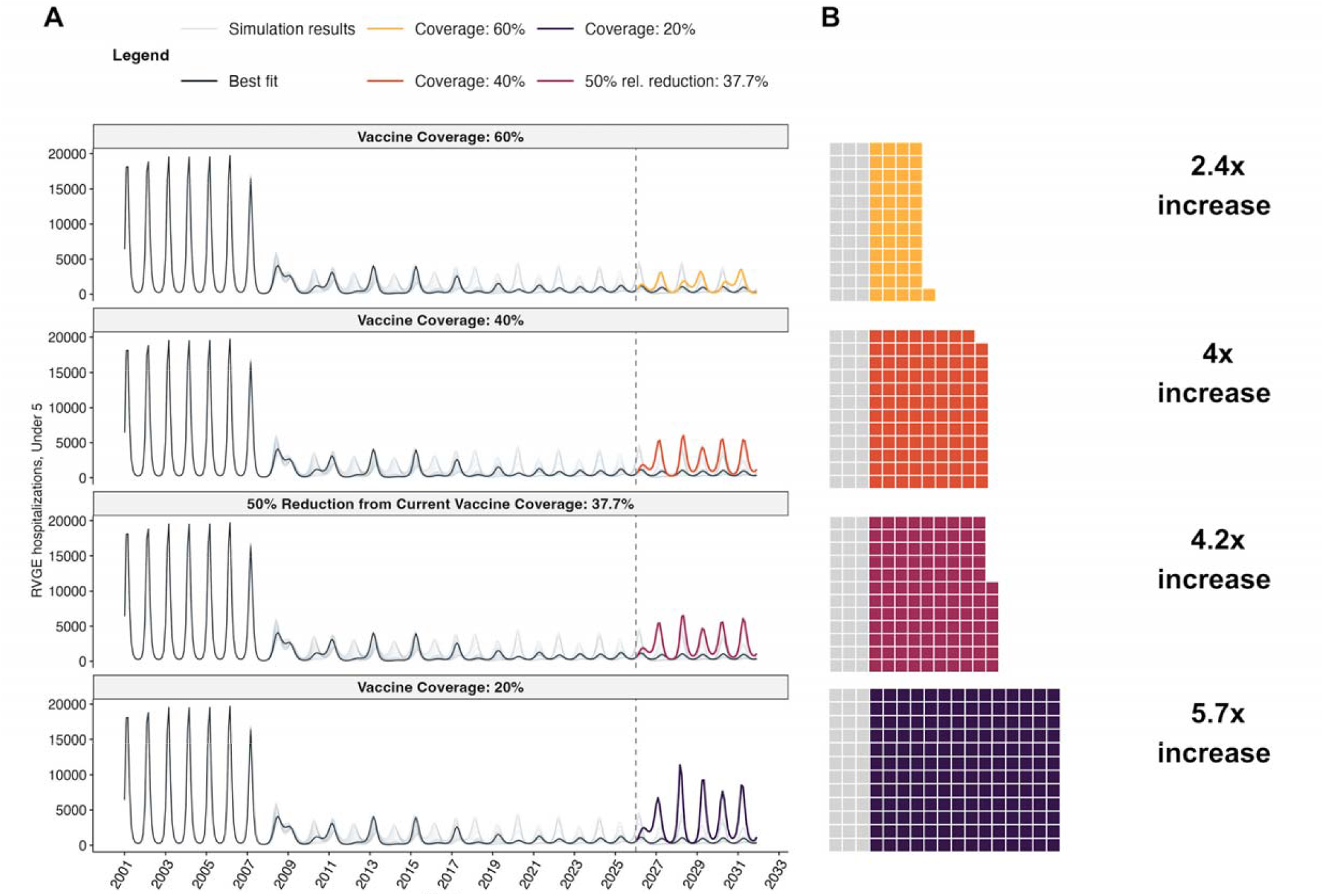
Projected impact of declining vaccine coverage on rotavirus gastroenteritis (RVGE) hospitalizations in the United States (US). (**A**) Time series of monthly RVGE hospitalizations from 2001 to 2031. Black lines represent model estimates based on vaccine coverage data for birth cohorts prior to 2021 and assuming coverage remains at current levels; the grey lines represent model estimates accounting for uncertainty in partial vaccine coverage and efficacy. Colored lines denote projections under three vaccine coverage scenarios: 60% coverage (yellow), 40% coverage (orange), 50% reduction from current coverage (37.7%; maroon), and 20% coverage (purple) beginning in July 2026. (**B**) Cumulative RVGE hospitalizations projected over five upcoming seasons (July 2026 through June 2031). Each square represents 1,000 hospitalizations. Grey squares indicate the baseline current vaccine coverage scenario. Colored squares represent additional hospitalizations at projected vaccine coverage scenarios.

Reflecting the diverse baseline of current rotavirus hospitalization rates and vaccine coverage levels across the US (**Figure 2A**), the relative increase in RVGE hospitalizations under these four vaccination scenarios varies greatly by state (**Figure 2B; Supplementary Figures 1-5**). In Massachusetts, where rotavirus vaccine coverage is high, a 50% relative decrease in coverage (from 94% to 47%) is estimated to lead to a 5.3-fold increase in the rate of RVGE hospitalizations (from 0.51 to 2.69 hospitalizations per 10,000 person-years) (**Figure 2**). While a biennial epidemic pattern is currently observed in Massachusetts, we predict that the state will return to annual outbreaks when coverage drops below 47% (**Supplementary Figure 1)**. In Mississippi, where rotavirus vaccine coverage is among the lowest in the nation, a 50% relative decrease to 29% coverage is projected to cause a 2-fold increase (from 7.31 to 14.62 hospitalizations per 10,000 person-years). Texas is predicted to experience the largest increase in RVGE hospitalizations (5.3-fold, from 7.36 to 39.34 hospitalizations per 10,000 person-years) if coverage decreases to 20%, likely due to the higher birth rate in Texas (**Figure 2B; Supplementary Table 3**).

**Figure 2.**
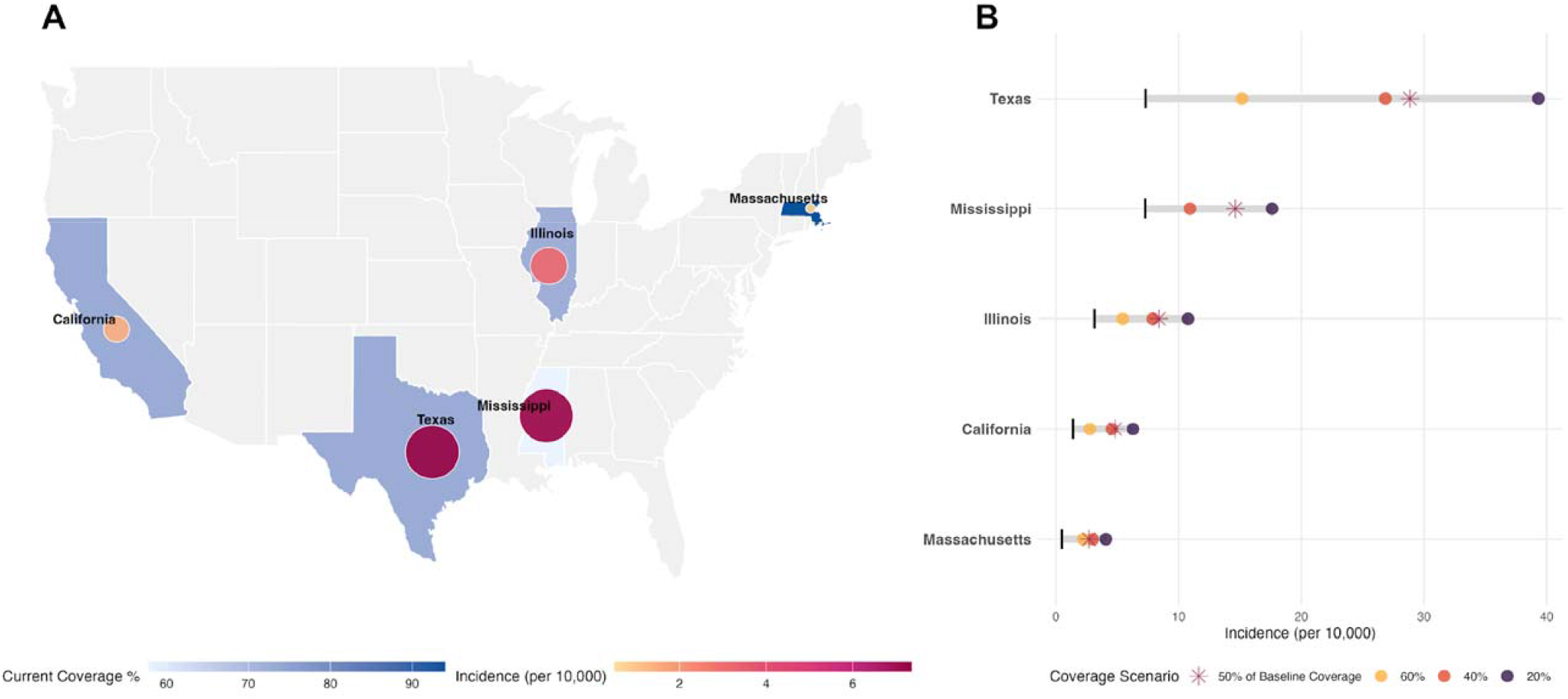
Geographic heterogeneity in rotavirus gastroenteritis (RVGE) hospitalization rates and projected changes under vaccine coverage shifts. **(A)** State-level baseline vaccination coverage and corresponding RVGE hospitalization rates for the July 2023-June 2025 period. Blue shading represents vaccination coverage, with darker tones indicating higher uptake. Superimposed yellow-to-red circles denote hospitalization rates; larger, darker circles represent higher baseline rates. **(B)** Projected state-level RVGE hospitalization rates across four vaccine coverage scenarios (60%, 40%, 20%, and a 50% relative reduction) for the 2027/28 and 2028/29 seasons (average for two seasons). Each dot represents a projected value for a specific coverage scenario; the asterisk (*) highlights the 50% relative coverage reduction scenario. Black vertical pipes (|) denote the baseline incidence for the 2023/24 and 2024/25 seasons, serving as a reference for projected shifts.

Post-licensure studies have found that rotavirus vaccination is associated with a small increase in the risk of intussusception following the first dose (1.5 [95% CI, 0.2 to 3.2] excess cases of intussusception per 100,000 vaccinees) (*15*). If rotavirus vaccine coverage were to decrease by 50% nationally, this would translate to about 1.5 million fewer infants vaccinated per year and potentially 22 fewer cases of vaccine-associated intussusception annually. Over the five-year period from July 2026 to June 2031, we predict that there will be 114,500 more RVGE hospitalizations if coverage were to drop by 50% nationally, which vastly exceeds the decrease in the number of possible vaccine-associated intussusception cases.

It is likely that the burden of the changes to rotavirus vaccine recommendations will fall disproportionately on states such as Texas and Mississippi that already experience higher rates of RVGE hospitalizations. In response to recent changes in CDC guidelines, several states, including Massachusetts and California, have announced that they will continue to follow the AAP-recommended vaccine schedule. Massachusetts currently has one of the highest rotavirus vaccine coverage rates in the nation and as a result experiences small, biennial epidemics of RVGE with a low hospitalization rate. If all states were to achieve the same vaccine coverage as Massachusetts, we predict an additional 4,600 RVGE hospitalizations could be prevented per year in the US in 2030/31 (**Supplementary Figure 6**).

Our analysis highlights the considerable increase in the burden of RVGE hospitalization that could result from downgrading the US rotavirus vaccine recommendation to “shared clinical decision-making”. The justification for changing this recommendation is not rooted in science, nor was the change rigorously reviewed by scientists and physicians according to standard evaluation frameworks, i.e., the Evidence to Recommendations framework used by Advisory Committee on Immunization Practices (*16*). Rotavirus is just one of several vaccines with proposed new, weaker recommendations for use at odds with those from AAP and endorsed by 12 medical groups representing 1 million health care professionals. This could signal the start of a new era of public health in America—an era in which diseases easily controlled by effective vaccines will once again sicken our children.

## Data Availability

Data and code used in this study are publicly available on GitHub: https://github.com/pitzerlab/Rotavirus-US

## Author contributions

Conceptualization: EOA, JK, MHC, SP, GSG, VEP

Methodology: EOA, VEP

Investigation: EOA, JK, VEP

Visualization: EOA, JK

Funding acquisition: VEP

Project administration: VEP

Supervision: GSG, VEP

Writing – original draft: MHC

Writing – review & editing: EOA, JK, SP, GSG, VEP

## Supplementary Materials

Materials and Methods

Fig. S1 to S6

Tables S1 to S4

