## Supplemental Figures S1 to S6; Supplemental Tables S1 to S4 for "Estimated Impacts of Rotavirus Vaccine Recommendation Changes in the U.S."

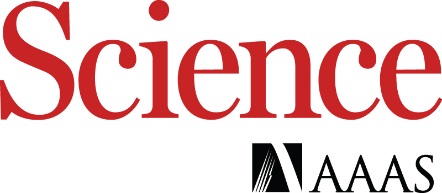

Supplementary Materials for

**Estimated Impacts of Rotavirus Vaccine Recommendation Changes in the U.S.**

Ernest O. Asare^1,2^, Jiye Kwon^1,2^, Melanie H. Chitwood^1,2^, Stephanie Perniciaro^1,2^, Gregg S. Gonsalves^1,2^, Virginia E. Pitzer^1,2^*

**The PDF file includes:**

Materials and Methods

Fig. S1 to S6

Tables S1 to S4

References

Materials and Methods

Rotavirus hospitalization data from Epic Cosmos

Data on rotavirus hospitalizations are from Epic Cosmos, a dataset created in collaboration with a community of Epic health systems representing more than 300 million patient records from over 1884 hospitals and 42,400 clinics from all 50 states, D.C., Lebanon, and Saudi Arabia. We counted the number of patients with hospitalized encounters containing a listing for a diagnosis of ICD-10 code A08.0 occurring among children <5 years old for residents of the United States (for national estimates of hospitalization rates) or for residents of Alabama, California, Illinois, Massachusetts, Mississippi, or Texas (respectively for state hospitalization rates) (Table S1). To calculate the denominator, we used the total number of patients <5 years old linked to any hospital visit over the 2023-2025 period, not accounting for multiple hospital visits per patient. To calculate the incidence rate per 10,000 people per year, we then multiplied by 10,000 people and divided by two years.

Rotavirus transmission model

Using a previously calibrated and validated age-stratified rotavirus transmission model (*1, 2*), we assessed the impact of current vaccination coverage and projected the potential increase in rotavirus-associated gastroenteritis (RVGE) hospitalizations across multiple U.S. states associated with a possible reduction in coverage under the U.S. government’s shift to a shared clinical decision-making (SCDM) recommendation.

The model has previously been described in detail (*1, 2*). Briefly, the model assumes that individuals are born with maternal immunity. Once maternal immunity wanes, they become fully susceptible and can experience multiple rotavirus infections over their lifetime, with both susceptibility and disease severity decreasing after the first and second infection, with subsequent infections assumed to be mild or asymptomatic. Following an infection, individuals acquire temporary immunity, which wanes after 9 months, and they become susceptible to reinfection at a reduced rate. Only first and second infections are assumed to result in moderate-to-severe RVGE, which could lead to hospitalization. Each vaccine dose was assumed to mimic a natural rotavirus infection, providing partial protection against reinfection among those who respond (i.e. seroconvert) to each dose. Full protection against moderate-to-severe RVGE is achieved after responding to two or more doses. Fixed parameter estimates were based on a seminal birth cohort study by Velazquez et al (*3*), as well as household transmission studies (*4*) and immunological (*5-8*) and viral shedding (*9*) data, as described in Table S2.

The force of infection is modeled as a function of the prevalence of first (*I*_1_), second (*I*_2_), and subsequent (*I_A_*) infections times a seasonally-varying transmission coefficient (*β*(*t*)):

*λ*(*t*) = *β*(t)(*I*_1_(*t*) + *ρ*_2_*I*_2_(*t*) + *ρ_A_I_A_*(*t*)),

We assume that second and subsequent infections are less infectious (by factors *ρ*_2_ and *ρ_A_*, respectively). Seasonality in the rate of transmission is modeled using a simple sinusoidal function, such that *β*(*t*) = *β*_0_(1 + *b*cos(2*π*(*t* – *φ*))), where *β*_0_ is the baseline transmission rate, *b* is the amplitude of seasonality, and *φ* is the seasonal offset parameter. Children <1 year of age, 1-year olds, and 2-year olds are assumed to be at increased risk of acquiring rotavirus infection (*c*_0_, *c*_1_, and *c*_2_, respectively), based on a comparison of different mixing assumptions (*1, 2*). Finally, we assumed that a proportion *h* of moderate-to-severe RVGE cases were hospitalized.

The reporting fraction *h* and the transmission parameters (*β*_0_, *a, φ, c*_0_, *c*_1_, *c*_2_) were previously estimated by fitting to age-specific hospitalization data from the State Inpatient Databases of the Healthcare Cost and Utilization Project (Table S3), assuming that the observed number of hospitalizations in each age group and month was Poisson-distributed with mean equal to the model-predicted incidence (*1, 2*). The model was to fitted pre-vaccination hospitalization data for 16 states in the U.S., including California, Illinois, and Massachusetts (*1*). Parameter estimates for Texas were based on models previously fitted to data on laboratory-confirmed rotavirus cases from the National Respiratory and Enteric Viral Surveillance System (NREVSS), while parameter estimates for Mississippi were assumed to be equal to the average parameter estimates for models fitted to NREVSS data from Alabama, Arkansas, Louisiana, and Tennessee.

Model calibration and validation

The vaccination component was subsequently incorporated in the model, and the model was shown to accurately predict out-of-sample data from the U.S. for 2006-2008 (*1*) and data from New York City for 2008-2016 (*1, 2*), providing important sources of model validation.

To project the number of rotavirus hospitalizations in each of the five states and the U.S. through 2032, we used rotavirus vaccination coverage estimates from the National Immunization Survey-Child (*10*) at both national and state levels. The most recent publicly available estimates are for the 2021 birth cohort with the full rotavirus series (two doses Rotarix or three doses of Rotateq) by 36 months of age (Table S4). In the absence of data on 1-dose or partial rotavirus vaccine series coverage, a parameter (*v*_1_) representing the relative coverage and response rate to the first rotavirus vaccine dose was calibrated to reproduce the estimated overall vaccine effectiveness estimate for 2006–2016 reported by Baker et al., who reported an overall reduction in the rate of rotavirus hospitalization of 78% (95% confidence interval: 71%, 83%) among children <5 years of age (*11*). We explored plausible ranges of *v*_1_ and retained parameter values that yielded a reduction in the model-predicted rate of hospitalization for 2007-2016 compared to pre-vaccination (2001-2005) within the reported 95% confidence interval for the same period, with the best-fit parameter estimate corresponding to the point estimate from Baker et al. (*11*).

Finally, given differences in the data sources for rotavirus hospitalizations, we adjusted the reporting fraction *h* by comparing predicted rotavirus hospitalization rates for July 2023 to June 2025 from the models with estimates derived from the Epic Cosmos database for the corresponding time period, both at the national level and across the five states. The reporting fraction was adjusted to ensure that these two rates were equal before projecting hospitalization rates through 2032.

Vaccine coverage scenarios

To evaluate the potential impact of reduced vaccination coverage under current policy recommendations, we simulated a range of alternative coverage scenarios, including a 50% relative reduction from current levels and fixed coverage levels of 60%, 40%, and 20%. We further assessed a scale-up scenario in which the United States and four states with lower baseline coverage achieve 94% coverage, corresponding to the level observed in Massachusetts, the state with the highest reported vaccination coverage. The impact of each scenario was quantified as the percentage change in disease burden from July 2026 to June 2031, relative to a baseline scenario based on current vaccination coverage.

Table S1.

Number of rotavirus hospitalizations from Epic Cosmos database, July 2023 to June 2025.

| Region | Hospitalizations | Person-Years | Rate (per 10,000 PY) |
| --- | --- | --- | --- |
| United States | 9,948 | 12,073,349 | 4.12 |
| California | 235 | 827,263 | 1.42 |
| Illinois | 331 | 516,647 | 3.20 |
| Massachusetts | 27 | 266,604 | 0.51 |
| Mississippi | 234 | 160,150 | 7.31 |
| Texas | 2,117 | 1,437,688 | 7.36 |

Table S2. Fixed model parameter values.

| Parameter | Symbol | Value | Source |
| --- | --- | --- | --- |
| Rate of waning maternal antibodies | *ω_0_* | 0.333 mo^-1^ | (*12*) |
| Rate of waning immunity following primary and secondary infection | *ω_1_* | 0.111 mo^-1^ | (*6*) |
| Rate of waning immunity following asymptomatic infection | *ω_2_* | 0.083 mo^-1^ | (*13*) |
| Rate of recovery from primary infection | *γ_1_* | 4.3 mo^-1^ | (*14*) |
| Rate of recovery from secondary and asymptomatic infection | *γ_2_* | 8.6 mo^-1^ | (*13, 15*) |
| Relative susceptibility following first infection | *σ_1_* | 0.62 | (*3*) |
| Relative susceptibility following second infection | *σ_2_* | 0.35 | (*3*) |
| Relative infectiousness of secondary infection | *ρ_1_* | 0.5 | (*4*) |
| Relative infectiousness of asymptomatic infection | *ρ_2_* | 0.1 | (*4*) |
| Proportion of first infections with severe diarrhea | *d_1_* | 0.11 | (*3*) |
| Proportion of second infections with severe diarrhea | *d_2_* | 0.029 | (*3*) |

Table S3. Fixed model parameters and population under age 5 for 2024 across five states and the United States.

| **Parameter** | **US** | **CA** | **IL** | **MA** | **MS** | **TX** |
| --- | --- | --- | --- | --- | --- | --- |
| Transmission rate (*R*_0_) | 23.25 | 20.594 | 30.029 | 25.087 | 27.376 | 27.376 |
| Amplitude of seasonality in transmission | 0.047 | 0.045 | 0.056 | 0.052 | 0.072 | 0.041 |
| Phase shift of seasonal transmission | 0.636 | 0.647 | 0.711 | 0.695 | 0.577 | 0.653 |
| Hospitalization fraction | 0.041 | 0.035 | 0.076 | 0.024 | 0.019 | 0.013 |
| Relative risk of infection for <1-year-olds | 1.555 | 1.582 | 0.971 | 1.419 | 1.159 | 1.159 |
| Relative risk of infection for 1 to <2-year-olds | 2.312 | 2.342 | 1.75 | 2.561 | 2.058 | 2.058 |
| Relative risk of infection for 2 to <3-year-olds | 1.738 | 1.77 | 1.31 | 1.78 | 1.462 | 1.462 |
| Crude birth rate (2024) | 10.65 | 10.19 | 9.89 | 9.55 | 11.50 | 12.52 |
| Under 5 population (2024) | 19,257,434 | 2,252,895 | 677,147 | 339,388 | 185,489 | 1,983,137 |

Table S4.

State-level vaccine uptake data from the National Immunization Survey-Child, 2021 birth cohort.

| Region | Vaccine Coverage (%) |
| --- | --- |
| United States | 75.4 |
| California | 73.0 |
| Illinois | 72.3 |
| Massachusetts | 94.0 |
| Mississippi | 57.8 |
| Texas | 78.9 |

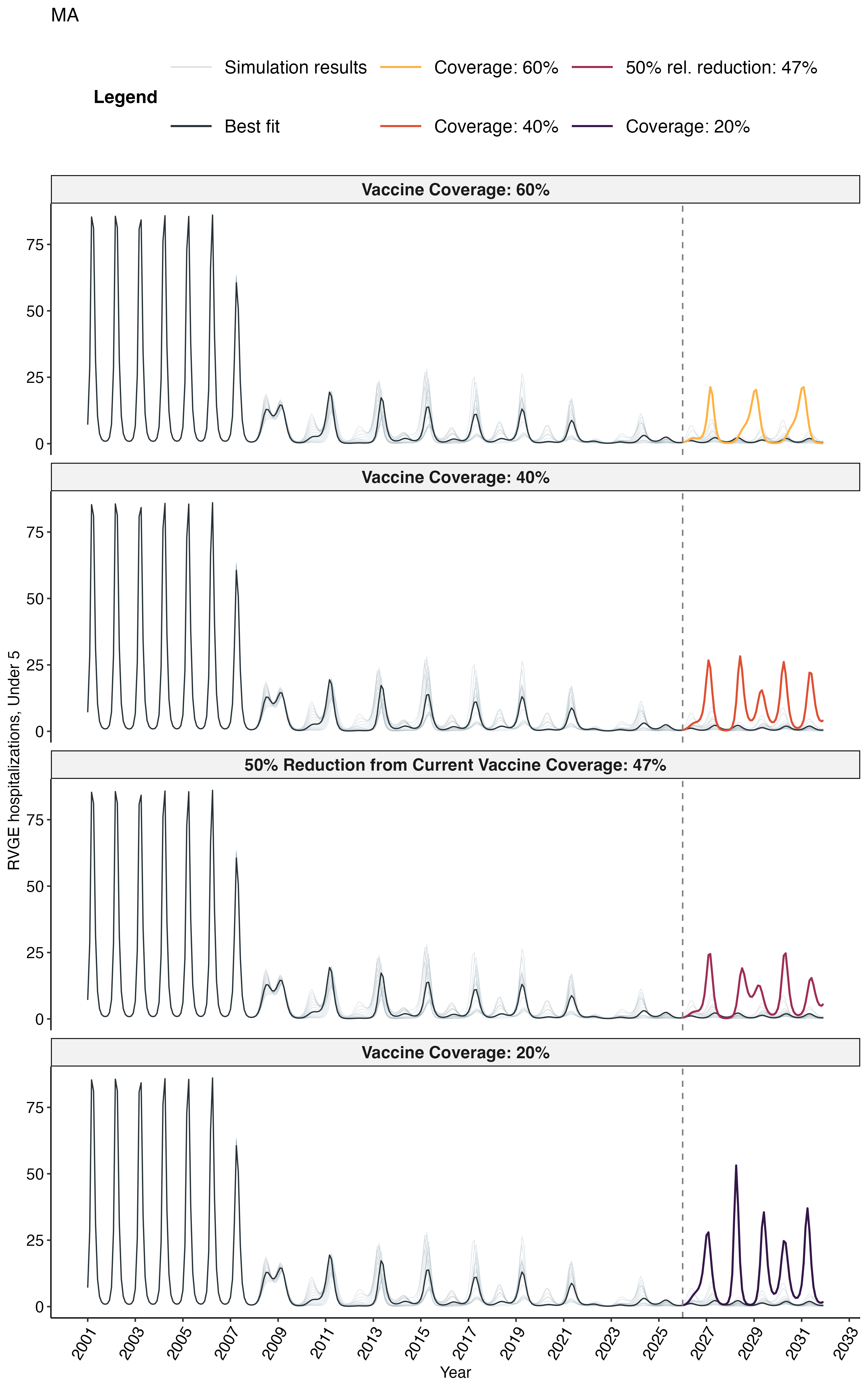

**Fig. S1. Projected impact of declining vaccine coverage on rotavirus gastroenteritis (RVGE) hospitalizations in Massachusetts.** Time series of monthly RVGE hospitalizations from 2001 to 2031. Black lines represent model estimates based on vaccine coverage data for birth cohorts prior to 2021 and assuming coverage remains at current levels; the grey lines represent model estimates accounting for uncertainty in partial vaccine coverage and efficacy. Colored lines denote projections under three vaccine coverage scenarios: 60% coverage (yellow), 40% coverage (orange), 50% reduction from current coverage (47%; maroon), and 20% coverage (purple) beginning in July 2026.

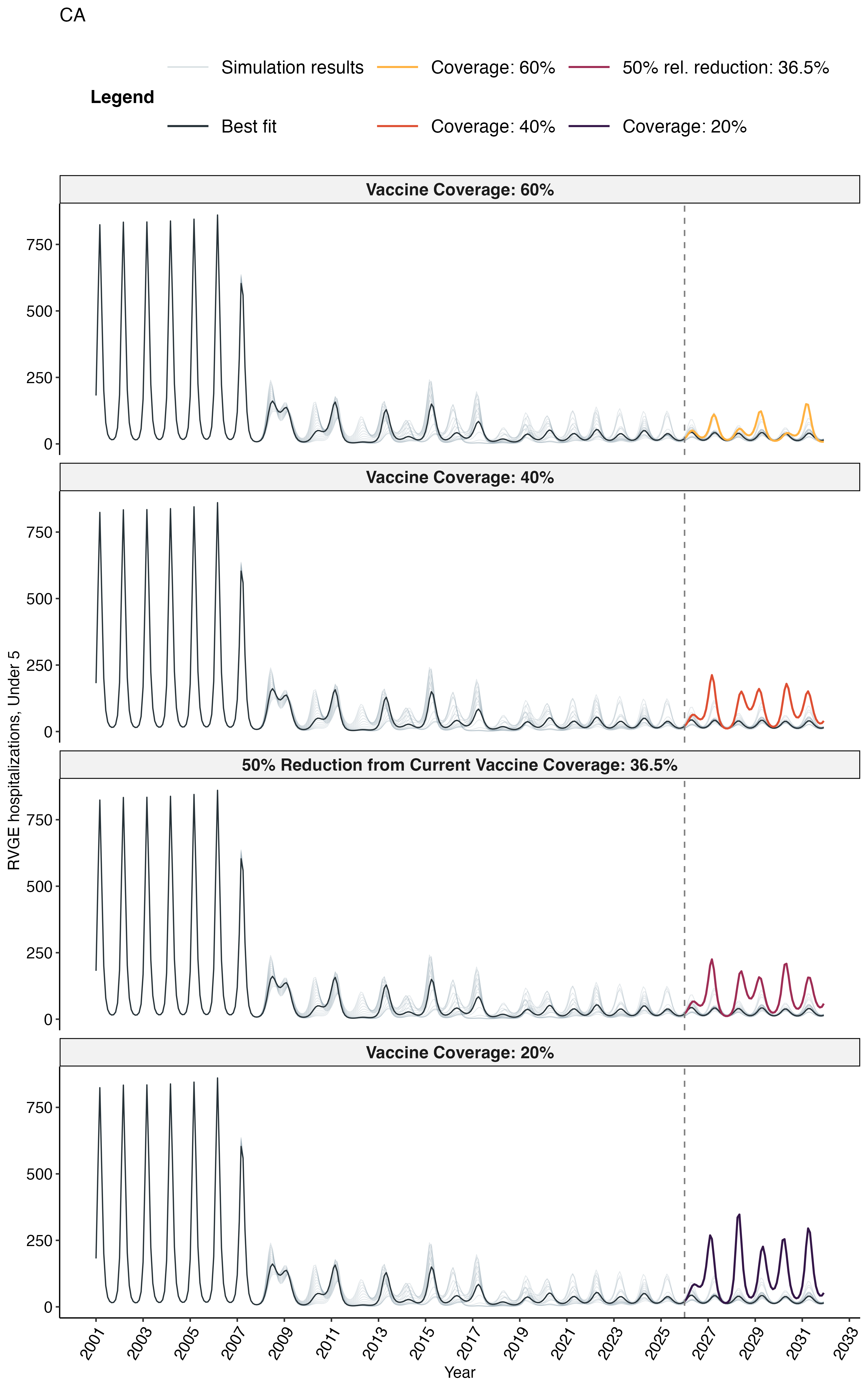

**Fig. S2. Projected impact of declining vaccine coverage on rotavirus gastroenteritis (RVGE) hospitalizations in California.** Time series of monthly RVGE hospitalizations from 2001 to 2031. Black lines represent model estimates based on vaccine coverage data for birth cohorts prior to 2021 and assuming coverage remains at current levels; the grey lines represent model estimates accounting for uncertainty in partial vaccine coverage and efficacy. Colored lines denote projections under three vaccine coverage scenarios: 60% coverage (yellow), 40% coverage (orange), 50% reduction from current coverage (36.5%; maroon), and 20% coverage (purple) beginning in July 2026.

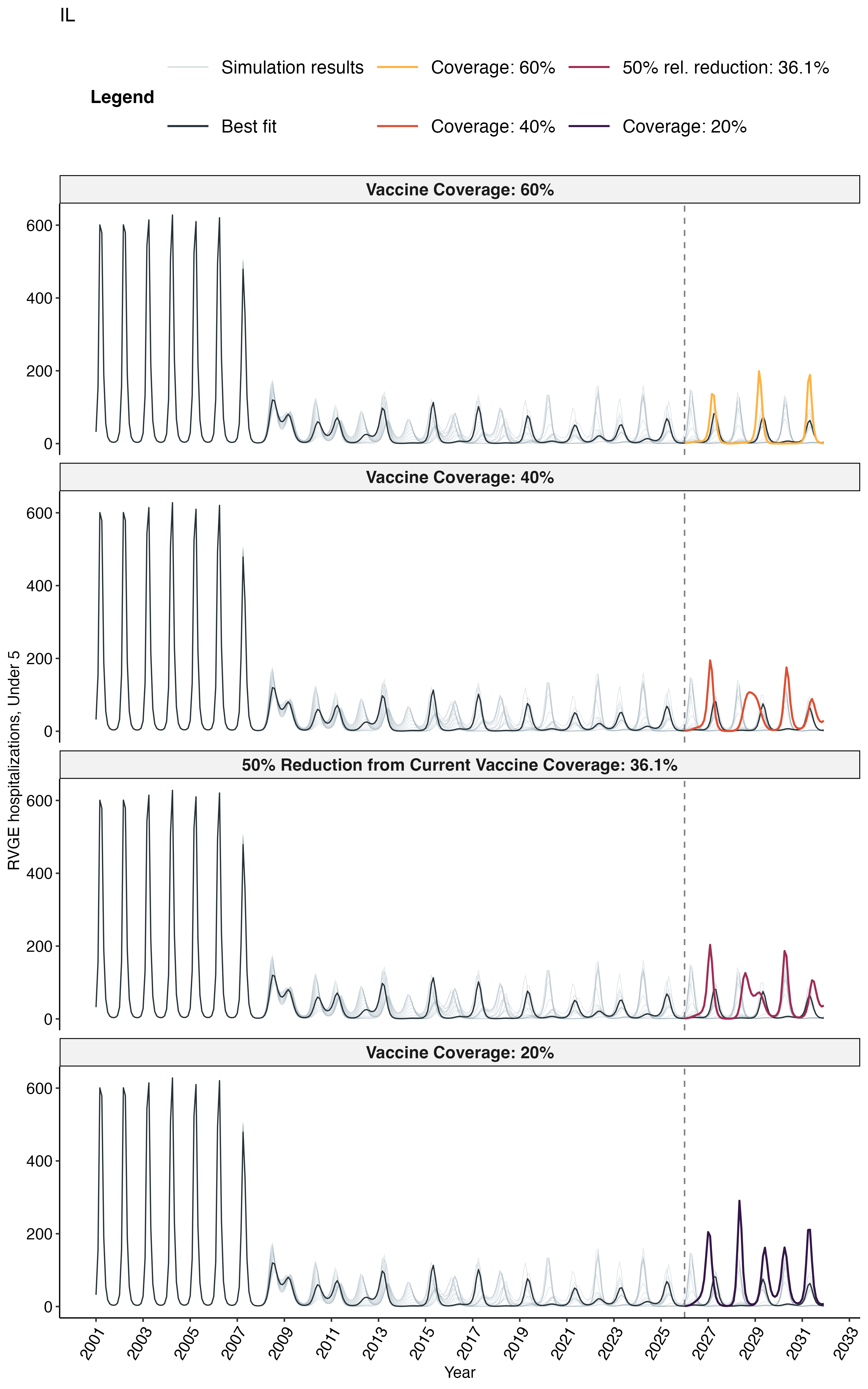

**Fig. S3. Projected impact of declining vaccine coverage on rotavirus gastroenteritis (RVGE) hospitalizations in Illinois.** Time series of monthly RVGE hospitalizations from 2001 to 2031. Black lines represent model estimates based on vaccine coverage data for birth cohorts prior to 2021 and assuming coverage remains at current levels; the grey lines represent model estimates accounting for uncertainty in partial vaccine coverage and efficacy. Colored lines denote projections under three vaccine coverage scenarios: 60% coverage (yellow), 40% coverage (orange), 50% reduction from current coverage (36.1%; maroon), and 20% coverage (purple) beginning in July 2026.

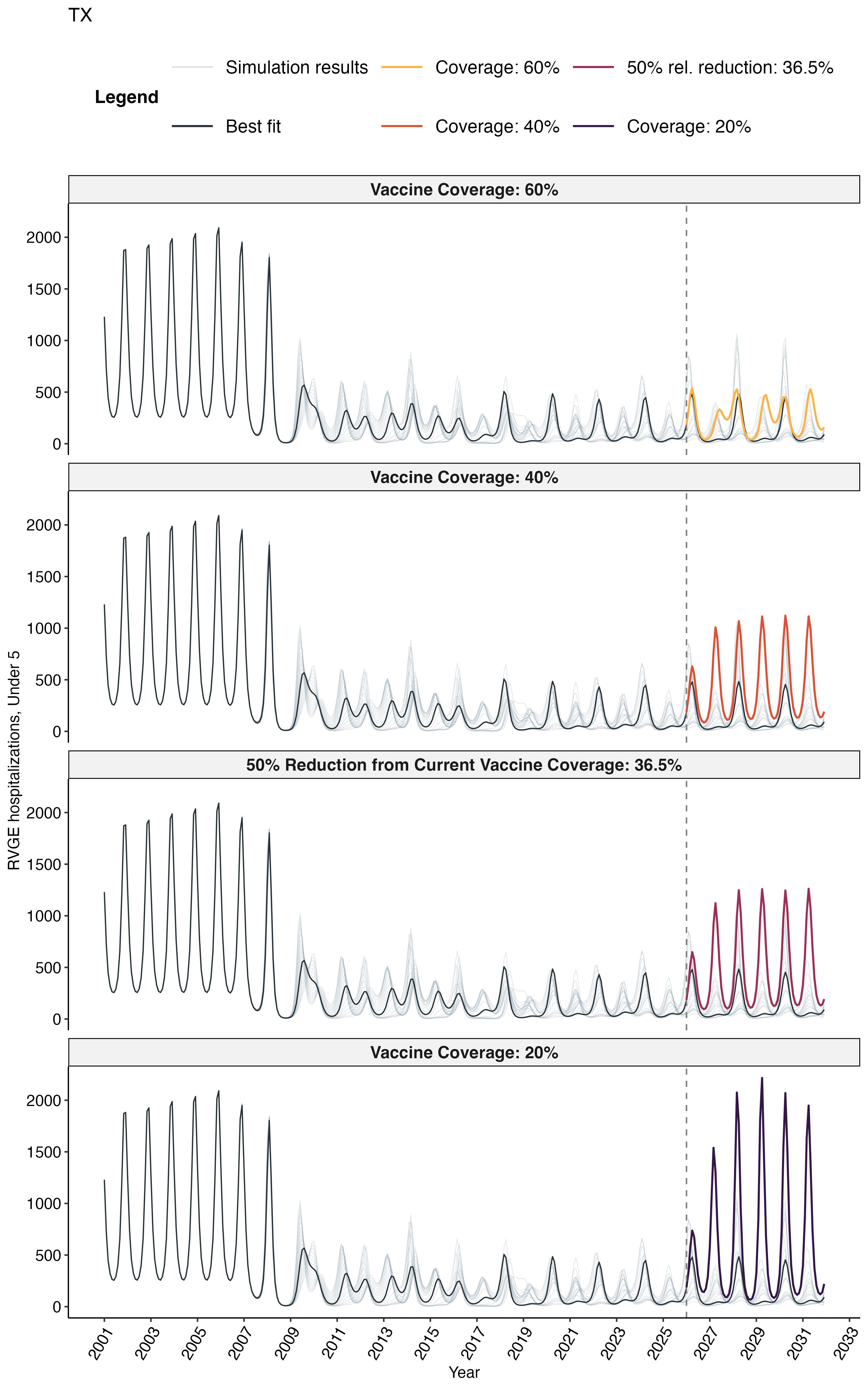

**Fig. S4. Projected impact of declining vaccine coverage on rotavirus gastroenteritis (RVGE) hospitalizations in Texas.** Time series of monthly RVGE hospitalizations from 2001 to 2031. Black lines represent model estimates based on vaccine coverage data for birth cohorts prior to 2021 and assuming coverage remains at current levels; the grey lines represent model estimates accounting for uncertainty in partial vaccine coverage and efficacy. Colored lines denote projections under three vaccine coverage scenarios: 60% coverage (yellow), 40% coverage (orange), 50% reduction from current coverage (36.5%; maroon), and 20% coverage (purple) beginning in July 2026.

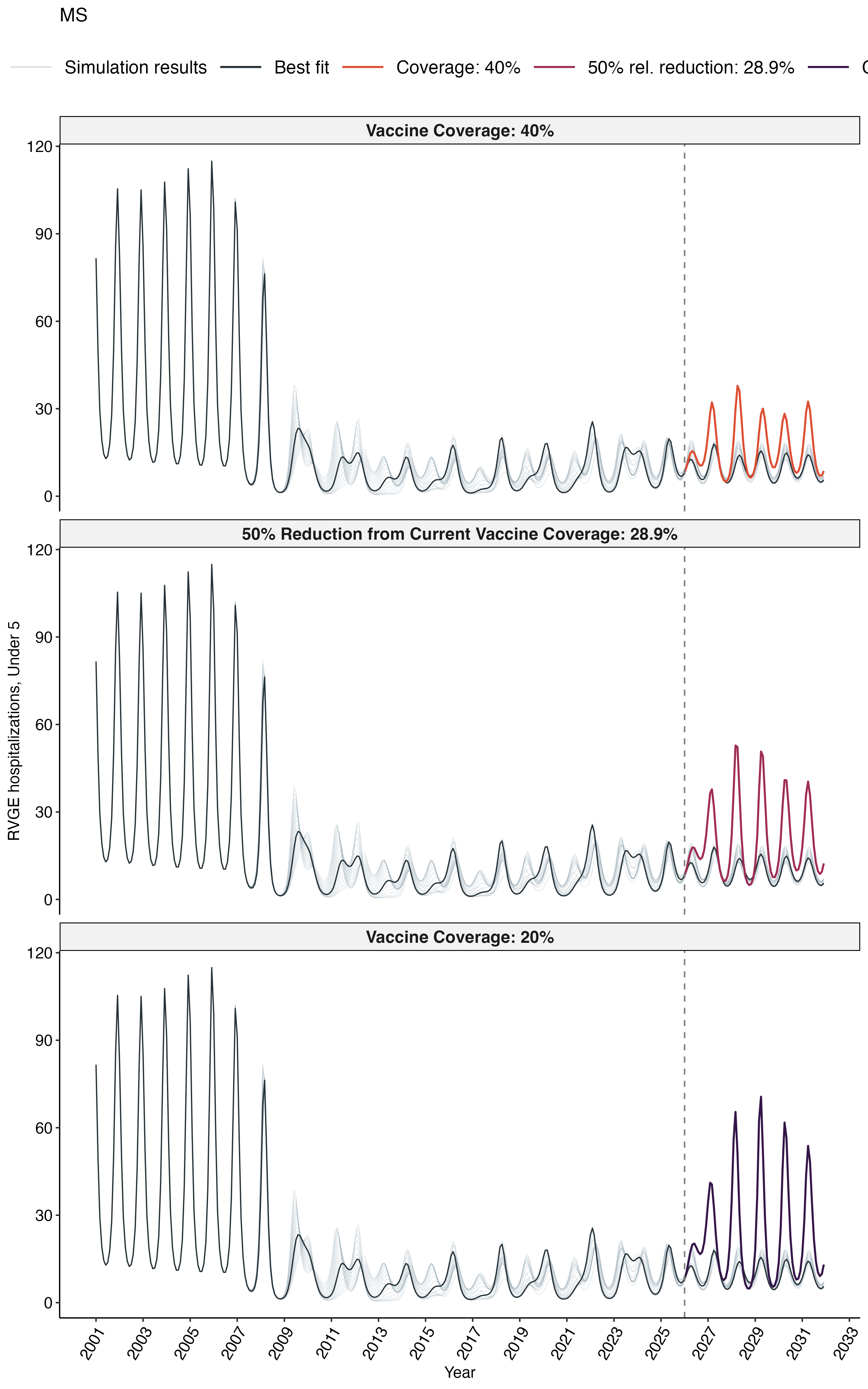

**Fig. S5. Projected impact of declining vaccine coverage on rotavirus gastroenteritis (RVGE) hospitalizations in Mississippi.** Time series of monthly RVGE hospitalizations from 2001 to 2031. Black lines represent model estimates based on vaccine coverage data for birth cohorts prior to 2021 and assuming coverage remains at current levels; the grey lines represent model estimates accounting for uncertainty in partial vaccine coverage and efficacy. Colored lines denote projections under three vaccine coverage scenarios: 40% coverage (orange), 50% reduction from current coverage (28.9%; maroon), and 20% coverage (purple) beginning in July 2026.

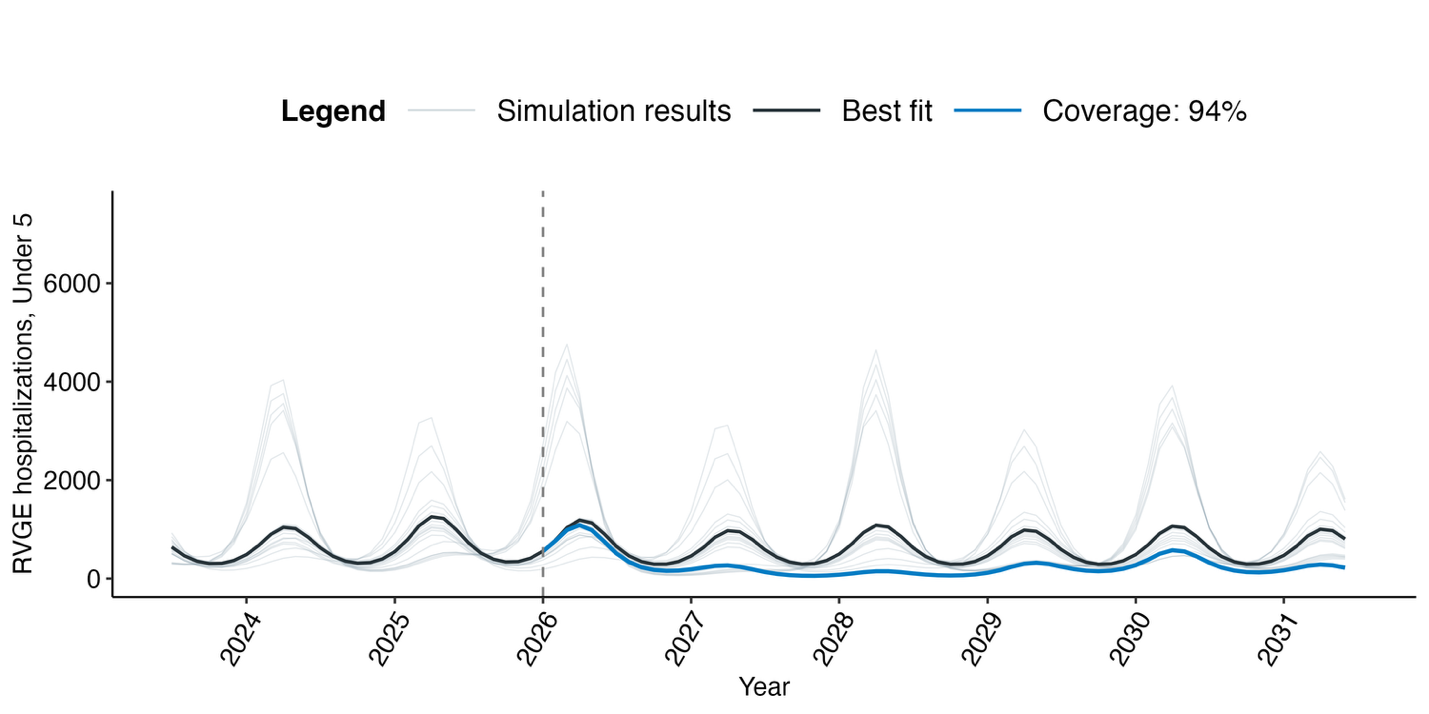
Fig. S6. Projected impact of increasing national vaccine coverage to 94% on rotavirus gastroenteritis (RVGE) hospitalizations in the United States. Time series of monthly RVGE hospitalizations from July 2023 through June 2031. Values prior to 2026 represent model estimates based on historical coverage data. Projections from July 2026 onward compare the status quo (black) against a scenario where overall vaccination coverage increases to 94% (blue). Thin background grey lines represent simulation results for different values of the relative effectiveness of the first vaccine dose (*v*_1_).
